# Multi-omic characterization reveals a distinct molecular landscape in young-onset pancreatic cancer

**DOI:** 10.1101/2023.03.28.23287894

**Authors:** Ifeanyichukwu Ogobuiro, Yasmine Baca, Jennifer R Ribeiro, Phillip Walker, Gregory C Wilson, Prateek Gulhati, John L Marshall, Rachna T Shroff, David Spetzler, Matthew J Oberley, Daniel E Abbott, Hong Jin Kim, David A Kooby, Shishir K Maithel, Syed A Ahmad, Nipun B. Merchant, Joanne Xiu, Peter J. Hosein, Jashodeep Datta

## Abstract

**Purpose:** Using a real-world database with matched genomic-transcriptomic molecular data, we sought to characterize the distinct molecular correlates underlying clinical differences between young-onset pancreatic cancer (YOPC; <50-yrs.) and average-onset pancreatic cancer (AOPC; ≥70-yrs.) patients.

**Methods:** We analyzed matched whole-transcriptome and DNA sequencing data from 2430 patient samples (YOPC, n=292; AOPC, n=2138) from the Caris Life Sciences database (Phoenix, AZ). Immune deconvolution was performed using the quanTIseq pipeline. Overall survival (OS) data was obtained from insurance claims (n=4928); Kaplan-Meier estimates were calculated for age-and molecularly-defined cohorts. Significance was determined as FDR-corrected *P*-values (*Q*)<0.05.

**Results:** YOPC patients had higher proportions of mismatch repair-deficient (dMMR)/microsatellite instability-high (MSI-H), *BRCA2*-mutant, and *PALB2*-mutant tumors compared with AOPC patients, but fewer *SMAD4-, RNF43-, CDKN2A-*, and *SF3B1-*mutant tumors. Notably, YOPC patients demonstrated significantly lower incidence of *KRAS* mutations compared with AOPC patients (81.3% vs. 90.9%; *Q*=0.004). In the *KRAS-*wildtype subset (n=227), YOPC tumors demonstrated fewer *TP53* mutations and were more likely driven by *NRG1* and *MET* fusions, while *BRAF* fusions were exclusively observed in AOPC patients. Immune deconvolution revealed significant enrichment of natural killer (NK) cells, CD8^+^ T cells, monocytes, and M2 macrophages in YOPC patients relative to AOPC patients, which corresponded with lower rates of *HLA-DPA1* homozygosity. There was an association with improved OS in YOPC patients compared with AOPC patients with *KRAS*-wildtype tumors (median 16.2 [YOPC-*KRAS*^WT^] vs. 10.6 [AOPC-*KRAS*^WT^] months; *P*=0.008) but not *KRAS*-mutant tumors (*P*=0.084).

**Conclusion:** In this large, real-world multi-omic characterization of age-stratified molecular differences in PDAC, YOPC is associated with a distinct molecular landscape that has prognostic and therapeutic implications.

## INTRODUCTION

Pancreatic ductal adenocarcinoma (PDAC) is a highly lethal malignancy with a 5-year survival rate of 12%,^1^ and is a leading cause of cancer-related mortality in the United States.^2^ PDAC is typically diagnosed in the seventh decade of life, referred to as average-onset pancreatic cancer (AOPC);^1,3^ however, young-onset pancreatic cancer (YOPC)—defined as diagnosis at less than 50 years of age^4,5^—constitutes 6-9% of newly detected PDAC and has steadily increased in incidence over the last two decades,^5-11^ as was recently highlighted by a new analysis of data from the National Program of Cancer Registries database.^12^ Emerging data indicates that smoking,^4,5^ alcohol use,^13^ obesity^14^, and family history^13,15,16^ are risk factors for YOPC. YOPC also skews toward male sex,^7,11^ although rates in women—particularly Black women—are rising faster than in men.^6,7,12^

The heterogeneity in the molecular landscape of PDAC that underpins its broad range of tumor phenotypes is one of the driving forces for suboptimal outcomes despite modern multimodal therapy.^17^ However, clinically annotated tumor profiling database studies such as the Know Your Tumor study have demonstrated that PDAC patients who received therapies matching actionable mutations had longer survival than those who received non-matched therapies.^18^ Moreover, The Cancer Genome Atlas (TCGA) analysis of PDAC revealed that, excluding *KRAS* and *CDKN2A*, 42% of patients could be candidates for molecularly informed clinical trials.^19^ The increasing armamentarium of precision medicine approaches for PDAC patients emphasizes the critical need to understand tumor-level molecular differences between YOPC and AOPC patients that might inform personalized therapy in this subset of patients.

Efforts to describe molecular differences between YOPC and AOPC have been hampered by a lack of real-world, large-scale matched genomic and transcriptomic data, leading to conflicting conclusions between studies. For instance, Raffenne and colleagues found no substantial differences in the mutational landscape between YOPC and AOPC patients,^3^ while others have identified higher *SMAD4* mutation rates, increased activation of the TGF-β pathway,^20^ and differential expression of *CDKN2A* and *FOXC2* in YOPC tumors compared to AOPC.^21^ Despite these differences, some unifying signals have emerged, particularly that YOPC patients harbor fewer oncogenic somatic *KRAS* mutations but more pathogenic germline mutations than AOPC patients.^16,19,20^ Further complicating our understanding of this question are the conflicting survival outcomes observed in these studies, with most indicating that YOPC patients have improved survival, ^9,11,22,23^ but others showing either shorter or no difference in survival compared with AOPC patients. ^3,5,10,15,20,24^ Together, these results illustrate gaps in our understanding of the genomic and transcriptomic correlates underlying clinical differences between YOPC and AOPC patients.

In the present study, using a cohort of 2430 sequenced tumors—including 292 YOPC— in a real-world multi-institutional dataset, we sought to characterize the distinct molecular landscape associated with YOPC compared with AOPC and better understand molecular correlates underlying the divergent clinical outcomes in YOPC patients.

## METHODS

### Patient Samples

2430 histologically-confirmed PDAC samples were identified in the Caris Life Sciences database (Phoenix, AZ) with matched DNA sequencing, whole transcriptome sequencing, and immunohistochemistry data. We stratified these specimens into YOPC (<50 years at diagnosis, n=292) and AOPC (≥70 years, n=2138). Among YOPC, 179 were from metastases and 113 from primary tumors; among AOPC, 1167 were from metastases and 967 from primary tumors.

### Next-Generation Sequencing (NGS)

Tumor enrichment was achieved using manual microdissection of nuclear fast red (NFR) stained formalin-fixed, paraffin-embedded (FFPE) sections that were marked for areas with at least 20% tumor content. NGS was performed on genomic DNA using the NextSeq or NovaSeq 6000 platforms (Illumina, Inc., San Diego, CA). For NextSeq sequenced tumors, a custom-designed SureSelect XT assay was used to enrich 592 whole-gene targets (Agilent Technologies, Santa Clara, CA). For NovaSeq sequenced tumors, a hybrid pull-down panel of baits designed to enrich for >700 clinically relevant genes at high coverage and read-depth was used, along with a separate panel designed to enrich for an additional >20,000 genes at lower depth. Genetic variants were detected with >99% confidence and were categorized by board-certified molecular geneticists as previously described.^25^ Tumor mutational burden (TMB)-high was defined as ≥10 mutations/Mb.

### Immunohistochemistry (IHC)

FFPE sections on glass slides were stained for PD-L1 (clone SP142, Spring Biosciences) using automated staining techniques, per the manufacturer’s instructions, and were optimized and validated per CLIA/CAP and ISO requirements. Staining was identified as positive if its intensity on the membrane of the tumor cells was ≥2+ (on a semiquantitative scale of 0–3: 0 no staining, 1+ weak staining, 2+ moderate staining, or 3+ strong staining) and the percentage of positively stained cells was >5%.

### Mismatch Repair Deficiency (dMMR)/Microsatellite Instability-High (MSI-H) Status

A combination of multiple test platforms was used to determine dMMR/MSI-H status of the tumors profiled, including fragment analysis (FA, Promega, Madison, WI), IHC (MLH1, M1 antibody; MSH2, G2191129 antibody; MSH6, 44 antibody; and PMS2, EPR3947 antibody [Ventana Medical Systems, Tucson AZ]) and NGS. The three platforms generated highly concordant results as previously reported, ^26^ and in the rare cases of discordant results, MSI/MMR status of the tumor was determined in the order of IHC, FA and NGS.

### Whole Transcriptome Sequencing (WTS)

FFPE sections on glass slides were stained with NFR, and areas with at least 10% tumor content were marked for manual microdissection and mRNA isolation. WTS was performed using the Illumina NovaSeq platform (Illumina, Inc., San Diego, CA) and Agilent SureSelect Human All Exon V7 bait panel (Agilent Technologies, Santa Clara, CA); transcripts per million (TPM) were reported. Gene fusions were detected using the ArcherDx fusion assay and Illumina MiSeq platform as previously described.^27^ Immune cell fraction was calculated by the quanTIseq pipeline^28^ using deconvolution of bulk transcriptomic data. Gene set enrichment analysis (GSEA) and Metascape pathway analysis were performed on WTS data.^29,30^ HLA genotyping was performed using arcasHLA, an *in-silico* tool that infers HLA genotypes from RNA sequencing data.^31^ If a single HLA genotype was detected, the specimen was classified as “homozygous”, which can occur due to parental homozygosity or HLA loss of heterozygosity (LOH).

### Statistical Analysis

Clinicodemographic features were compared by Chi-square test, with *P*<0.05 considered statistically significant. Comparative analysis of molecular alterations in the cohorts were analyzed using Chi-square or Fisher exact tests. Tumor microenvironment cell fractions were analyzed among cohorts using non-parametric Kruskal-Wallis testing. Because these closely related cohorts are only differentiated by age, *P*-values<0.05 were highlighted as relevant trends. For a more stringent analysis of the differences between AOPC and YOPC, *P*-values were further corrected for multiple hypothesis testing using Benjamini-Hochberg method to avoid type I error, and adjusted *Q<*0.05 was considered statistically significant.

### Clinical Outcomes Data

Real-world overall survival (OS) information was obtained from insurance claims data and calculated from date of tissue collection to last contact. Kaplan-Meier estimates were calculated for YOPC and AOPC in the entire cohort of patients with clinical data in the Caris CODEai™ clinico-genomic database, which increased from the time of the molecular analysis (N=4928), and stratified by *KRAS* mutation status, which was available for 3116 patients (*KRAS*^WT^, N=393; *KRAS*^MUT^, N=2723). Significance was determined as log-rank *P*<0.05.

### Compliance Statement

This study was approved by the Institutional Review Board at the University of Miami and conducted in accordance with guidelines of the Declaration of Helsinki, Belmont report, and U.S. Common rule. Per 45 CFR 46.101(b)(4), this study utilized retrospective, de-identified clinical data, and no patient consent was necessary from the subjects.

### Data availability

Data presented in this study are not publicly available due to data size and patient privacy but are available on reasonable request from the corresponding author.

## RESULTS

### Clinicodemographic characteristics

At the time of molecular analysis, 2430 patient samples were molecularly annotated with genomic and transcriptomic data. A total of 4928 patients had available clinical outcomes data in the most recent query of the Caris CODEai clinico-genomic database, from which Kaplan-Meier curves were generated. Of the 2430 patients with molecular data, 292 patients (12%) were YOPC, with the median age in this YOPC cohort being 46 (IQR 41-48). Among 2138 AOPC patients (88%) included, the median age was 75 (IQR 72-79). There was a significant preponderance of male patients (65% vs. 52%, *P*<0.05) and current smokers (95% vs. 91%, *P*=0.023) in YOPC compared with AOPC patients, respectively (**Table 1**).

**Table 1.** Clinicodemographic characteristics of patients with pancreatic ductal adenocarcinoma (PDAC) from the Caris Life Sciences database. Specimens were stratified by young-onset pancreatic cancer (YOPC) and average-onset pancreatic cancer (AOPC) and analyzed by sex and smoking status. *P*-values were determined by Fisher exact test.

|  | YOPC (%) | AOPC (%) | *P*-value |
| --- | --- | --- | --- |
| Median age | 46 | 75 |  |
| **Sex** |  |  | **<0.05** |
| Males | 190 (65) | 1116 (52) |  |
| Females | 102 (35) | 1022 (48) |  |
| **Smoking status** |  |  | **0.023** |
| Current | 18 (95) | 142 (91) |  |
| Non-smoker | 1 (5) | 14 (9) |  |

### Comparative molecular landscape of YOPC and AOPC

Previous studies have reported differing prevalence of molecular alterations^3,19,20,32,33^ and a preponderance of germline mutations dominated by *BRCA1/2* and MMR genes in YOPC compared to AOPC patients.^16^ However, direct comparisons between YOPC and AOPC are scarce and have utilized smaller cohorts^3,20^. We analyzed clinically relevant and putative oncogenic tumor-related mutations and copy number alterations (CNA) in tumors derived from YOPC and AOPC patients in this large real-world cohort (**Supplementary Table S1**).

*KRAS* mutations were the most prevalent somatic alterations in both YOPC and AOPC (81.3% and 90.9%), followed by *TP53* (69.3% and 74.7%), *CDKN2A* (19.3% and 24.8%), and *SMAD4* (14.7% and 20.1%; **Fig. 1A-B**), respectively. Although germline mutational data were not available, YOPC patients had significantly higher rates of *somatic* alterations in homologous recombination deficiency-associated genes, specifically *BRCA2* (4.7% vs. 2.1%; *P*=0.008) and *PALB2* (1.4% vs 0.5%; *P*=0.044), compared with AOPC patients. YOPC patients were also noted to have higher rates of mismatch repair-deficient (dMMR)/microsatellite instability-high (MSI-H) tumors (2.8% vs. 0.8%; *P*=0.001) compared with AOPC patients.

**Figure 1.**
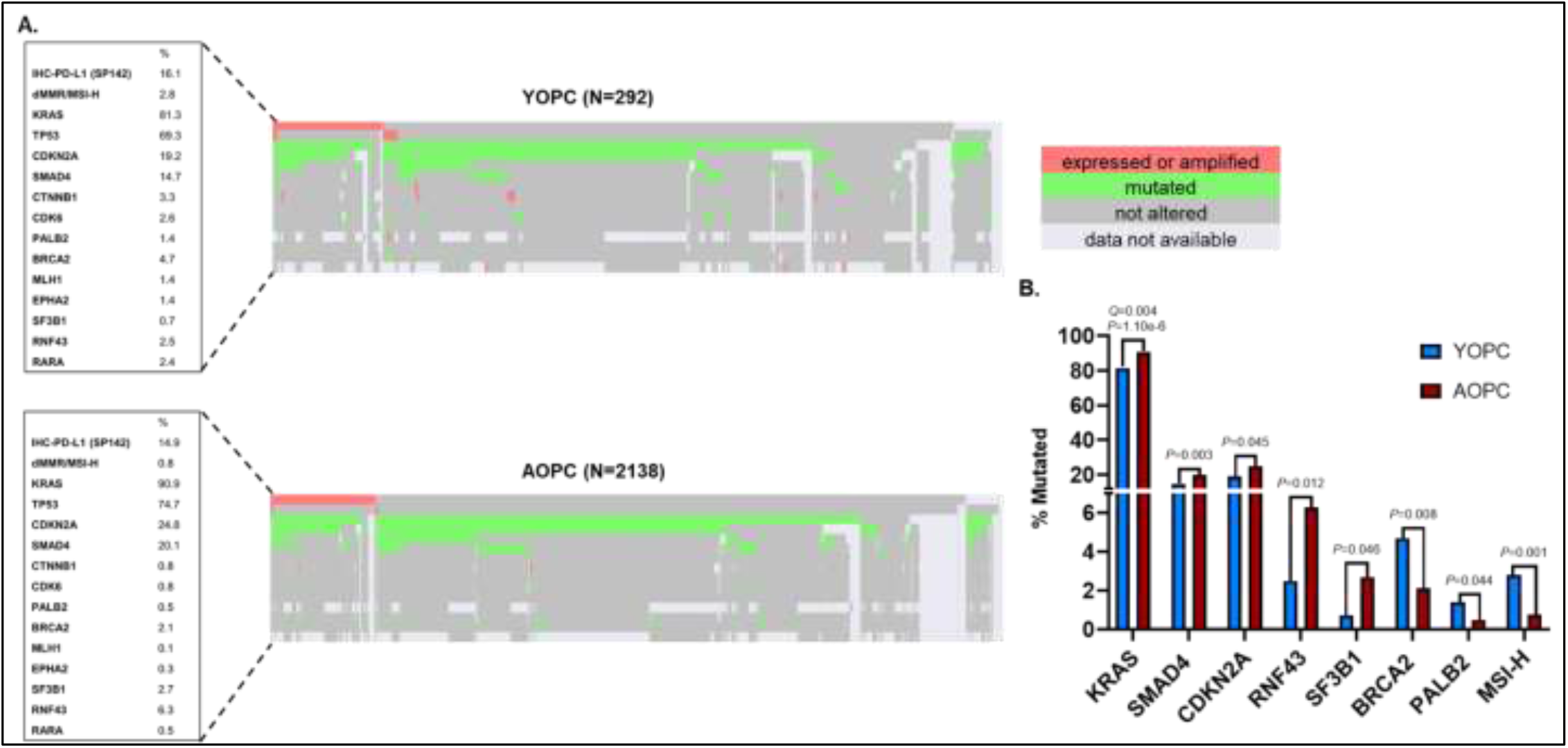
Molecular landscape of YOPC and AOPC. **(A)** Oncoprints displaying the pathogenic molecular alteration pattern of YOPC (N=292) and AOPC (N=2138) tumors. Columns represent tumor samples. Rows represent individual molecular biomarkers, whose percentages in the cohort are described in the boxes to the left of oncoprints. Pink, expressed or amplified; green, mutated; grey; not altered; light gray, data not available. **(B)** Bar graph showing statistically significant differential molecular alterations in YOPC (blue bars, N=292) versus AOPC (red bars, N=2138). *P*-values (Chi-square or Fisher exact tests) and FDR-adjusted *Q*-values are indicated above the compared groups for each molecular alteration.

Conversely, AOPC patients had significantly higher rates of oncogenic *KRAS* mutations compared with YOPC patients (90.9% vs 81.3%; *P*=1.10e-6; *Q*=0.004), as well as significantly higher rates of alterations in *CDKN2A* (24.8% vs 19.25%, *P*=0.045), *SMAD4* (20.1% vs. 14.7%; *P*=0.033), *RNF43* (6.3% vs. 2.5%; *P*=0.012), and *SF3B1* (2.7 % vs. 0.7%; *P*=0.046; **Fig. 1A-B**).

### Spectrum of alterations within KRAS^WT^ tumors in YOPC and AOPC patients

We next dissected the landscape of molecular alterations *within* the *KRAS*-wildtype (*KRAS*^WT^; N=227 [10.7%]) cohort given the significant enrichment of *KRAS*^WT^ tumors in YOPC patients (**Fig. 2A**). Previous studies have implicated the enrichment of mutations in *BRAF, CTNNB1*, and alternative RAS pathway genes^19^ in *KRAS*^WT^ PDAC. As such, we observed trends toward increased rates of *CTNNB1* mutations (17.7% vs. 4.0%; *P*=0.002) and reduced rates of oncogenic *TP53* mutations (21.3% vs. 44.4%; *P*=0.004) in YOPC-*KRAS*^WT^ tumors compared with AOPC-*KRAS*^WT^ tumors. Moreover, YOPC-*KRAS*^WT^ patients demonstrated higher rates of *MET* (4.1% vs. 0.6%; *P*=0.12) and *NRG1* (6.1% vs. 1.1%; *P*=0.07) fusions compared to AOPC-*KRAS*^WT^ patients, whereas *BRAF* fusions were exclusively concentrated in AOPC-*KRAS*^WT^ compared with YOPC-*KRAS*^WT^ tumors (6.8% vs. 0.0%; *P*=0.07; **Fig. 2B**). These results indicate distinct molecular vulnerabilities in *KRAS*^WT^ tumors when stratifying by age of PDAC onset.

**Figure 2.**
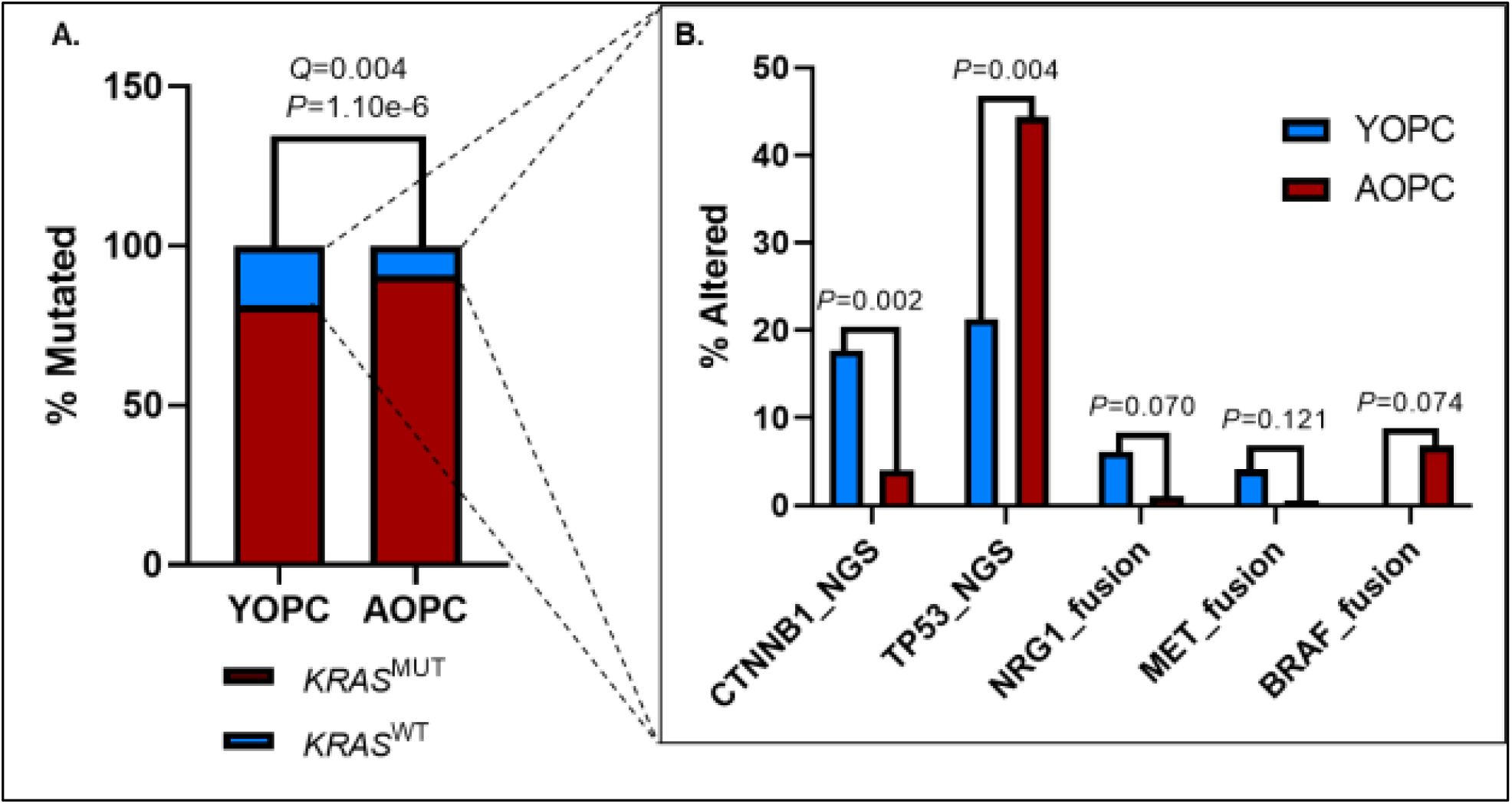
Spectrum of alterations within *KRAS-*wildtype YOPC and AOPC tumors. **(A)** Frequency of *KRAS*-wildtype *(KRAS*^WT^; blue, N=227) and *KRAS*-mutant (*KRAS*^MUT^; red, N=1970) tumors in YOPC and AOPC, determined by next-generation sequencing for pathogenic alterations. **(B)** *KRAS*^WT^ tumors, indicated in blue in **(A)**, were analyzed separately for differences in pathogenic molecular alterations. The spectrum of top mutations and fusions within *KRAS*^WT^ YOPC tumors (blue bars, N=49-51 [two YOPC-*KRAS*^WT^ patients lacked WTS data for fusions]) and AOPC tumors (red bars, N=176) is shown, with *P*-values (Chi-square or Fisher exact tests) indicated.

### Differentially regulated signaling pathways in tumor transcriptomes from YOPC vs. AOPC patients

To better understand how these genomic differences between YOPC and AOPC tumors influence downstream oncogenic and tumor microenvironment signaling, we performed GSEA comparing whole tumor transcriptomes in YOPC versus AOPC tumors. A relatively narrow number—i.e., total of 20—genes were significantly differentially expressed (*P*<0.05; *Q*<0.25) between YOPC and AOPC tumors (**Fig. 3A; Supplementary Table S2**). The top genes more highly expressed in YOPC tumors included carboxypeptidase B (*CPB2*), plasminogen (*PLG*), prothrombin (*F2*), and genes for fibrinogen alpha and beta chains (*FGA/FGB*), while plasminogen activator inhibitor 2 (*SERPINB2*) and interferon gamma (*IFNG*) had significantly higher expression in AOPC tumors. We then employed a less stringent *P*-value cutoff (P<0.25) in Metascape pathway analysis to clarify the transcriptomic nuances of these age-stratified PDAC cohorts. This analysis revealed that YOPC tumor transcriptomes were significantly enriched in pathways related to blood clotting cascade, extracellular matrix, cancer pathways, cytokine/inflammatory response, and angiogenesis (**Fig. 3B**).

**Figure 3.**
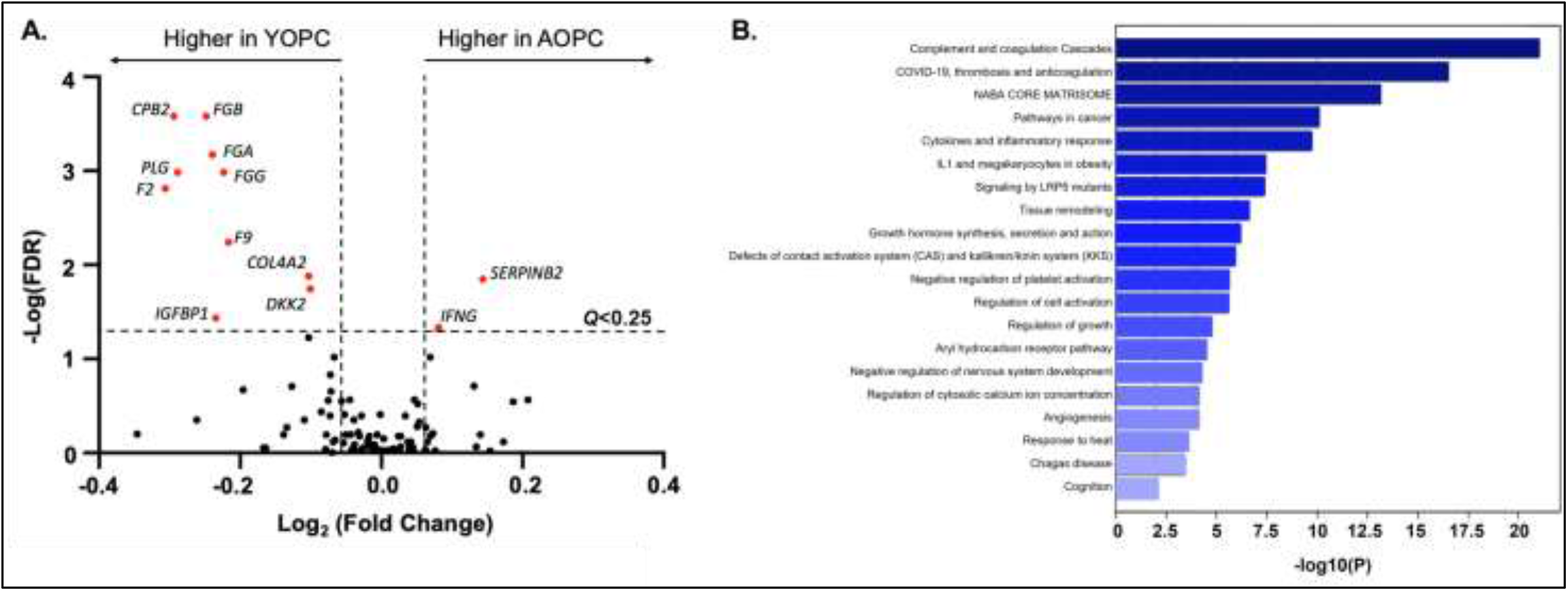
Differentially regulated signaling pathways in YOPC and AOPC. **(A)** The volcano plot shows differentially expressed genes (DEGs) between YOPC (N=284) and AOPC (N=2089), with a cutoff of FDR-adjusted *Q*<0.25. Genes to the left indicated in red are significantly higher in YOPC, while genes to the right indicated in red are significantly higher in AOPC. **(B)** Metascape pathway enrichment analysis was performed on 40 DEGs between YOPC and AOPC (*P*<0.25). The bar graph indicates canonical signaling pathways and biologic processes differentially enriched in the tumor transcriptomes of YOPC compared with AOPC tumor samples. The x-axis indicates statistical significance (-log10 *P*-value).

### Intra-tumoral immune deconvolution and HLA landscape in YOPC vs. AOPC tumors

While checkpoint immunotherapy in unselected patients has been decidedly unsuccessful in PDAC,^34-36^ some evidence points to possible success in highly selected patient groups.^37^ Studies showing the association between increased intratumoral T-cell infiltration and prolonged survival also suggest a role for the tumor immune microenvironment in dictating PDAC outcomes.^38-40^ Because we noted enrichment of select pathways and somatic alterations with diverse immunologic repercussions in YOPC tumors, we sought to determine differences in the tumor immune microenvironment between YOPC and AOPC using quanTIseq immune deconvolution of bulk tumor transcriptomes.^28^ While there were no significant differences in rates of TMB-high tumors, PD-L1 positivity (via IHC), or immune checkpoint gene expression between the cohorts (**Supplementary Fig. S1**), there was a statistically significant enrichment in computationally inferred signatures for natural killer (NK) cells (*P*=0.009; *Q*=0.039), CD8^+^ T-cells (*P*=0.043; *Q*=0.117), M2 macrophages (*P*=0.011; *Q*=0.039), and monocytic cells (*P*=0.002; *Q*=0.021) in YOPC compared to AOPC tumors (**Fig. 4A-B**).

**Figure 4.**
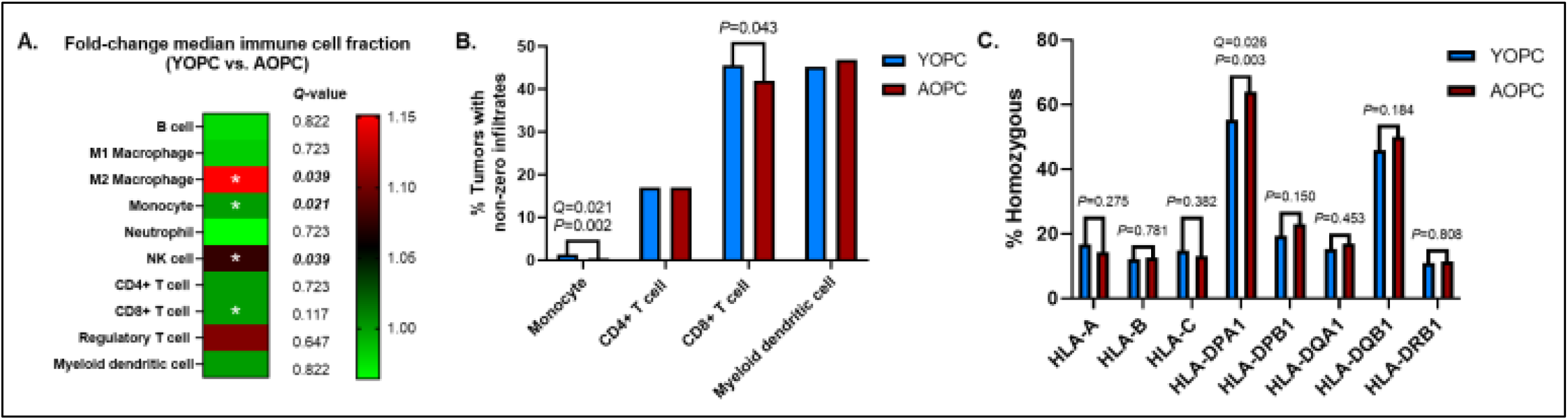
Intra-tumoral immune populations and HLA landscape in YOPC and AOPC. **(A)** Computationally inferred intra-tumoral Immune population between YOPC (N=284) and AOPC (N=2089). The heatmap indicates fold-change (YOPC vs AOPC) in median immune fraction according to quanTIseq. *P*-values were determined by non-parametric Kruskal-Wallis test. Asterisks indicate *P*<0.05, with *Q*-values shown to the right. **(B)** For cell types with median values of “0” (i.e., monocytes, CD4+ T cells, CD8+ T cells, and myeloid dendritic cells), the percentage of tumors with non-zero immune infiltrates were compared. **(C)** Differences in HLA landscape inferred from WTS data in YOPC (blue bars, N=284) compared to AOPC (red bars, N=2089). *P*-values (Chi-square or Fisher exact test) and FDR-adjusted *Q*-values are indicated above compared groups for each HLA gene.

To further understand potential major histocompatibility complex (MHC) determinants that might underlie these immunologic differences between cohorts, we examined HLA type and locus-specific expression inferred from RNA-sequencing data. We observed a significantly decreased rate of homozygosity in *HLA-DPA1* in YOPC compared to AOPC tumors (55.2% vs. 64.1%; *P*=0.003; *Q*=0.026; **Fig. 4C)**. Taken together, these associative data illustrate potential differences in immunogenicity related to cell-autonomous and/or non-autonomous mediators in YOPC tumors that might be contributory to differences in clinicopathologic outcomes between YOPC and AOPC patients.

### Overall survival of YOPC and AOPC patients stratified by KRAS^MUT^ and KRAS^WT^

The Caris CODEai dataset had 4928 patients with insurance claims-related follow-up/death information for analysis, but limited clinicopathologic data precluding stage- and treatment-stratified comparisons between YOPC and AOPC cohorts. Notwithstanding, we observed significantly longer OS in YOPC compared to AOPC patients (14.9 vs. 10.8 months; *P*<0.00001; **Fig. 5A**). Given the differences in frequency of *KRAS*-altered tumors between cohorts, we further stratified this analysis to determine if *KRAS* alteration status impacts OS. *KRAS* mutation data was available for 3116 PDAC patients with clinical information in the CODEai dataset. YOPC-*KRAS*^WT^ patients had significantly prolonged OS compared with AOPC-*KRAS*^WT^ patients (16.2 vs. 10.6 months; *P*=0.008; **Fig. 5B**). However, there was no difference in OS between YOPC and AOPC patients with *KRAS*^MUT^ tumors (12.9 vs. 10.0 months; *P*=0.084; **Fig. 5C**). These data suggest that survival differences between YOPC and AOPC patients in the overall cohort may be driven by survival variation specifically in patients harboring *KRAS*^WT^ tumors.

**Figure 5.**
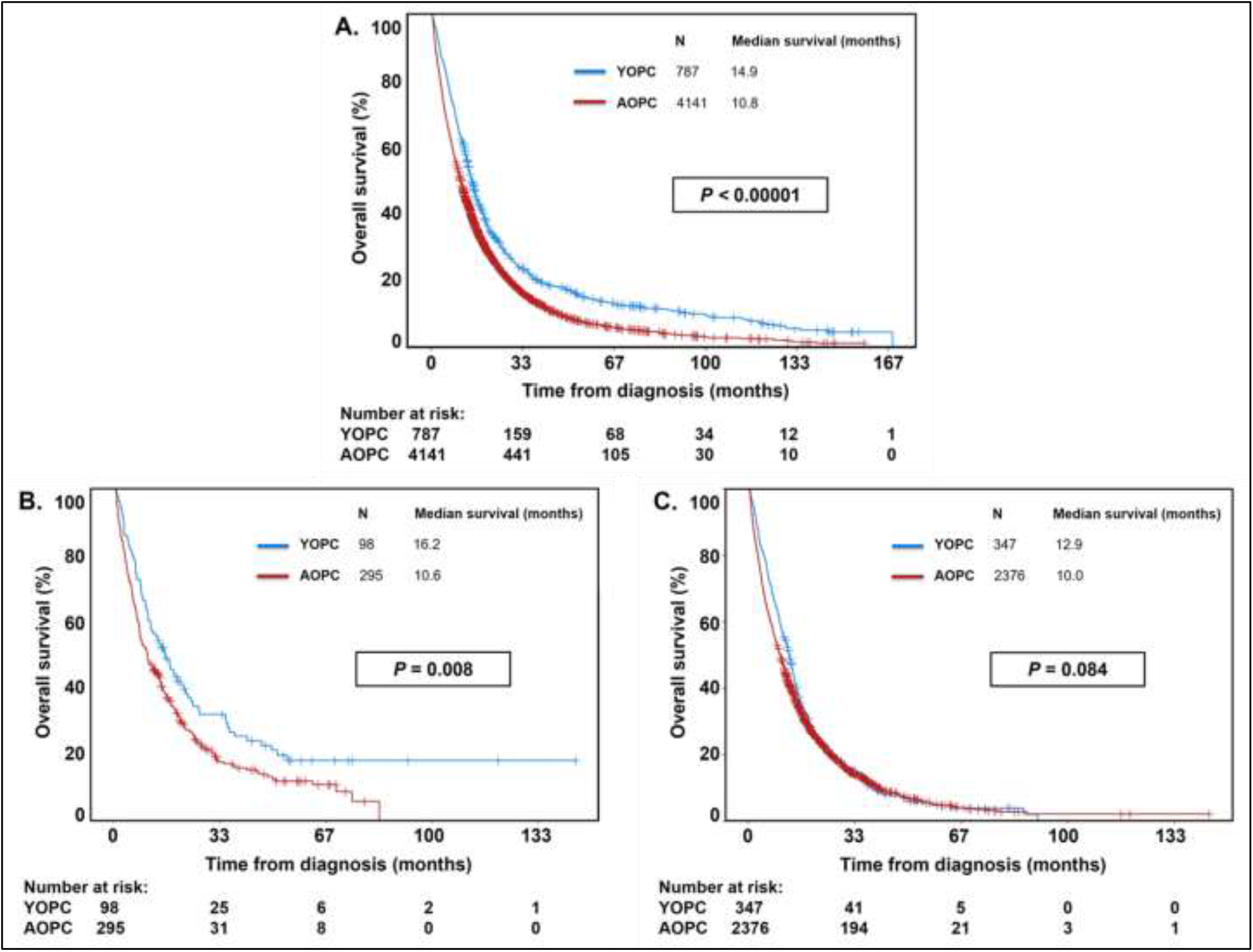
Overall survival (OS) of YOPC and AOPC patients stratified by *KRAS*^MUT^ and *KRAS*^WT^. **(A)** Kaplan-Meier curves depict the OS of YOPC patients (blue line, N=787) versus AOPC patients (red line, N=2753) in the entire PDAC cohort with clinical outcome data (N=4141 total). **(B-C)** All PDAC cases with *KRAS* mutation data available were stratified by *KRAS* status. Kaplan-Meier curves depict the OS of YOPC (blue line, N=98) versus AOPC (red line, N=295) patients with *KRAS*^WT^ tumors (N=393 total; **B**) and YOPC (blue line, N=347) and AOPC (red line, N=2376) patients with *KRAS*^MUT^ tumors (N=2723 total; **C**).

## DISCUSSION

The present study represents the largest pragmatic molecular comparison of YOPC versus AOPC. Our data reinforce previously observed epidemiologic distinctions between YOPC and AOPC patients,^4,5,7,11^ specifically its male preponderance and association with active smoking behaviors in YOPC patients, as well as conclusively reveal a higher incidence of *KRAS*^WT^ tumors in YOPC. Within this *KRAS*^WT^ subset, we uncovered distinct molecular vulnerabilities when stratifying by age—i.e., *MET* and *NRG1* fusions in YOPC-*KRAS*^WT^ and *BRAF* fusions in AOPC-*KRAS*^WT^. Among the unstratified cohort, YOPC patients demonstrated higher rates of somatic alterations in homologous recombination deficiency genes, higher prevalence of dMMR/MSI-H, and enrichment of NK cells and T cells. Finally, our data reconcile conflicting prior evidence by demonstrating improved survival in YOPC compared with AOPC patients, which may not only reflect the reduced prevalence of the virulent oncogenic drivers *KRAS, SMAD4*, and *CDKN2A* in YOPC tumor genomes, but may also be driven by the significantly longer survival of YOPC-*KRAS*^WT^ versus AOPC-*KRAS*^WT^ patients.

While the success of targeted and immune-based therapies has significantly lagged in PDAC compared with other solid tumors, the Know Your Tumor study conducted by the Pancreatic Cancer Action Network illustrated the oncologic importance of molecularly matched targeted therapies in advanced PDAC patients ^18^. To that end, our data provide a biologic map of the distinct molecular vulnerabilities in YOPC patients that might be exploited therapeutically. For instance, while *KRAS* mutations (with their rapidly evolving therapeutic landscape^41,42^) are ubiquitous in the broader cohort, our data reveal novel age-restricted molecular alterations in *KRAS*^WT^ tumors that may be clinically actionable; *NRG1, MET*, and *BRAF* fusions each have associated targeted therapies (e.g., afatinib, capmatinib, and encorafenib/vemurafenib, respectively).^43-45^ Moreover, given recent data indicating the benefit of polyadenosine diphosphate (ADP)-ribose polymerase (PARP) inhibitors in patients with germline or somatic mutations in homologous recombination deficiency genes,^46^ the enrichment of somatic *BRCA2* and *PALB2* alterations in YOPC tumors suggests that a higher proportion of YOPC patients may ultimately be eligible for therapies targeting the DNA damage response pathway. Taken together, these data call for heightened awareness among clinician and non-clinician stakeholders of the distinct genomic landscape in YOPC patients and underscore the importance of routine NGS testing in younger patients presenting with newly diagnosed advanced PDAC to inform potential molecularly targeted therapeutic approaches in these patients.

Exploration of the transcriptomes differentially expressed between YOPC and AOPC revealed enrichment of pathways associated with thrombotic cascades, extracellular matrix, cancer pathways, and cytokine/inflammatory response. This differentially expressed transcriptome in YOPC patients in the current study, however, is not strongly consistent with previous—albeit underpowered—studies that revealed enrichment in pathways predominantly related to hedgehog signaling and hypoxia in YOPC.^3,20^ This lack of concordance might be attributable to our substantially larger cohort size and/or the inherent heterogeneity of patients enrolled in this pragmatic “real world” study capturing data with wide geographic and clinicodemographic variability, which present novel insights into the genotype-immunophenotype chasm in YOPC.^47^ Multiple pathways enriched in YOPC tumors converged upon regulation of the tumor immune microenvironment with various predicted effects. For instance, cytokine signaling, cancer pathways, and angiogenic signaling may restrict tumor immunity.^48-50^ Conversely, the significant reduction in *HLA-DPA1* homozygosity—which has been previously associated with dampened antigen presentation and checkpoint blockade efficacy^51^—and associated increases in computationally inferred adaptive immune sub-populations (i.e., NK and CD8^+^ T cells) in YOPC suggest a less immunosuppressive and more immunostimulatory microenvironment. Altogether, these findings underscore the need for deeper investigation and functional characterization of cell-autonomous and non-autonomous immunologic repercussions in YOPC tumors.

Our study has several limitations. First, while the classification of YOPC and AOPC into <50 and ≥70-year age cutoffs was informed by prior studies,^3-5^ this arbitrary distinction may underestimate subtle molecular differences in YOPC patients. Second, while several of the reported genomic differences did not achieve significance by multiple hypothesis testing, we felt it important to report these novel signals with the recognition that our study compares molecular determinants in two closely related PDAC patient populations differentiated *solely* by a 20-year age gap. As such, molecular features distinguishing such cohorts are undoubtedly subtle and further validation in larger cohorts are warranted. Third, the lack of clinical annotation (e.g., performance status, resection status, stage, BMI, and multimodality treatment information) in the Caris CODEai dataset precluded meaningful stage- and treatment-stratified comparisons between cohorts or inclusion of multivariable analyses to account for confounding by these clinical parameters.

Given the rise in young-onset PDAC diagnosis in recent years, ^5-11^ these data are a timely addition to an expanding compendium of molecular taxonomy that highlights the clinical and phenotypic heterogeneity observed in this distinct cohort of patients.^4,5,7,11,16,19-21^ Furthermore, novel genomic and transcriptomic signals observed in YOPC tumors may offer a putative molecular basis for the divergent clinical outcomes observed in this population. Moving forward, these data could be incorporated into future trial design to allow more precise selection and stratification of YOPC and AOPC patients for elements of multimodality and/or novel therapies, with the goal of improving contemporary survival outcomes in this lethal malignancy.

## Data Availability

All data produced in the present study are available from the Precision Oncology Alliance of Caris Life Sciences upon reasonable request

