## Supplementary Materials for "Multi-omic characterization reveals a distinct molecular landscape in young-onset pancreatic cancer"

### SUPPLEMENTARY INFORMATION

| Supplementary Table S1. Molecular alterations in YOPC and AOPC. | | | | | | | | | |
|---|---|---|---|---|---|---|---|---|---|
| **Pathway** | **Biomarker (mutation, CNA, or fusion)** | **Positive (AOPC)** | **Negative (AOPC)** | **Percentage (AOPC)** | **Positive (YOPC)** | **Negative (YOPC)** | **Percentage (YOPC)** | ***P*-value** | ***Q*-value** |
| **RTK/RAS** | NGS-KRAS | 1749 | 176 | 90.9% | 221 | 51 | 81.3% | ***1.10E-06*** | ***0.004*** |
| | CNA-EGFR | 1 | 1914 | 0.1% | 2 | 268 | 0.7% | 0.004 | 1 |
| | NGS-RET | 0 | 1951 | 0.0% | 1 | 276 | 0.4% | 0.008 | 1 |
| **TGF-β** | CNA-SMAD2 | 0 | 1893 | 0.0% | 1 | 266 | 0.4% | 0.008 | 1 |
| | NGS-SMAD4 | 391 | 1553 | 20.1% | 40 | 233 | 14.7% | 0.033 | 1 |
| **WNT** | NGS-CTNNB1 | 16 | 1934 | 0.8% | 9 | 267 | 3.3% | 0.0003 | 0.704 |
| | NGS-RNF43 | 123 | 1832 | 6.3% | 7 | 270 | 2.5% | 0.012 | 1 |
| **Cell cycle** | CNA-CDK6 | 16 | 1881 | 0.8% | 7 | 261 | 2.6% | 0.008 | 1 |
| | NGS-CDKN2A | 472 | 1435 | 24.8% | 52 | 219 | 19.2% | 0.045 | 1 |
| **Chromatin remodeling** | NGS-SMARCB1 | 2 | 1953 | 0.1% | 3 | 275 | 1.1% | 0.001 | 0.992 |
| | CNA-SMARCB1 | 1 | 1916 | 0.1% | 2 | 269 | 0.7% | 0.004 | 1 |
| **Hedgehog** | NGS-SMO | 0 | 1952 | 0.0% | 1 | 276 | 0.4% | 0.008 | 1 |
| **Homologous Recombination** | NGS-BRCA2 | 41 | 1894 | 2.1% | 13 | 262 | 4.7% | 0.008 | 1 |
| | NGS-PALB2 | 9 | 1943 | 0.5% | 4 | 272 | 1.4% | 0.044 | 1 |
| **MMR** | NGS-MLH1 | 2 | 1949 | 0.1% | 4 | 272 | 1.4% | 5.34E-05 | 0.185 |
| **mRNA splicing** | NGS-SF3B1 | 53 | 1885 | 2.7% | 2 | 272 | 0.7% | 0.046 | 1 |
| **PI3K** | NGS-TSC1 | 0 | 1952 | 0.0% | 2 | 275 | 0.7% | 0.0002 | 0.484 |
| | NGS-MTOR | 0 | 1947 | 0.0% | 1 | 275 | 0.4% | 0.008 | 1 |
| | CNA-PIK3R2 | 0 | 791 | 0.0% | 1 | 126 | 0.8% | 0.013 | 1 |
| **Other** | CNA-KIF5B | 0 | 1827 | 0.0% | 2 | 258 | 0.8% | 0.0002 | 0.492 |
| | NGS-JAK1 | 2 | 1925 | 0.1% | 3 | 271 | 1.1% | 0.001 | 0.992 |
| | NGS-TMEM127 | 0 | 1133 | 0.0% | 1 | 143 | 0.7% | 0.005 | 1 |
| | CNA-TRIM27 | 0 | 1912 | 0.0% | 1 | 268 | 0.4% | 0.008 | 1 |
| | CNA-EPHA3 | 0 | 1870 | 0.0% | 1 | 263 | 0.4% | 0.008 | 1 |
| | CNA-IL7R | 0 | 1908 | 0.0% | 1 | 269 | 0.4% | 0.008 | 1 |
| | CNA-RALGDS | 0 | 679 | 0.0% | 1 | 101 | 1.0% | 0.010 | 1 |
| | CNA-MLLT1 | 0 | 773 | 0.0% | 1 | 124 | 0.8% | 0.013 | 1 |
| | CNA-ELL | 0 | 763 | 0.0% | 1 | 123 | 0.8% | 0.013 | 1 |
| | NGS-PRKAR1A | 2 | 1938 | 0.1% | 2 | 272 | 0.7% | 0.022 | 1 |
| | CNA-RARA | 4 | 794 | 0.5% | 3 | 124 | 2.4% | 0.025 | 1 |
| | CNA-PRDM16 | 2 | 736 | 0.3% | 2 | 112 | 1.8% | 0.031 | 1 |
| | NGS-EPHA2 | 3 | 1098 | 0.3% | 2 | 136 | 1.4% | 0.040 | 1 |

**Supplementary Table S1. Molecular alterations in YOPC and AOPC.** Numbers of positive and negative cases and overall percentages for top pathogenic gene mutations, fusions, and copy number alterations (CNA) are shown. All genes shown displayed differences with *P*<0.05 (Chi-square or Fisher exact test). FDR-corrected *Q*<0.05 is bolded and italicized.

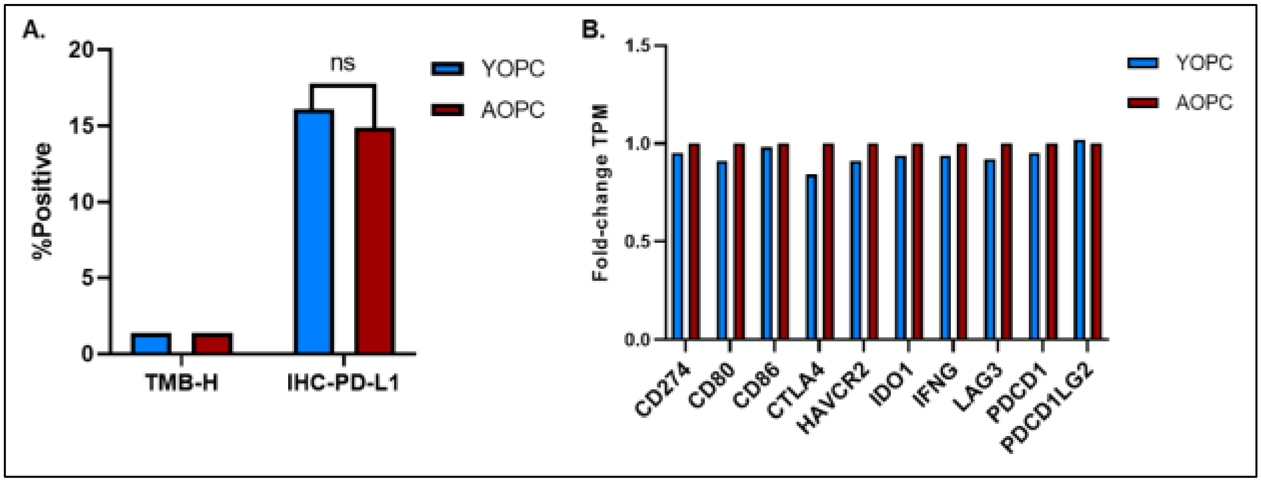


**Supplementary Figure S1. Immune landscape of YOPC and AOPC. (A)** Percentage of cases positive for immunotherapy biomarkers TMB-H and PD-L1(+) for YOPC (blue bars, N=273-292 [a small number of patients lacked PD-L1 IHC or dMMR/MSI-H analysis]) and AOPC (red bars, N=2028-2138 [a small number of patients lacked PD-L1 IHC or dMMR/MSI-H data]). Statistical analysis was performed by Chi-square or Fisher exact test. **(B)** Fold-change gene expression levels in transcripts per million (TPM) of immune checkpoint genes between YOPC (blue bars, N=284) and AOPC (red bars, N=2089). AOPC expression is set to 1 for each gene. There were no statistically significant differences in immune gene expression between YOPC and APOC (determined by Mann-Whitney U test).

| Supplementary Table S2. List of differentially expressed genes between YOPC and AOPC. | | | | | | |
|---|---|---|---|---|---|---|
| **Gene** | **age>=70** | **age<70** | **p-value** | **fold change** | **log2 fold change** | **q-value** |
| FGB | 213.0639 | 253.103 | 5.35E-06 | 0.841807 | -0.24844 | 0.000262 |
| CPB2 | 10.73488 | 13.16266 | 4.15E-06 | 0.815556 | -0.29415 | 0.000262 |
| FGA | 180.9031 | 213.5293 | 2.06E-05 | 0.847205 | -0.23922 | 0.000673 |
| PLG | 31.45159 | 38.41664 | 5.28E-05 | 0.818697 | -0.2886 | 0.001035 |
| FGG | 241.6043 | 282.0574 | 4.47E-05 | 0.856579 | -0.22334 | 0.001035 |
| F2 | 9.288775 | 11.4822 | 9.46E-05 | 0.808971 | -0.30584 | 0.001545 |
| F9 | 5.103183 | 5.930754 | 0.000409 | 0.860461 | -0.21682 | 0.005722 |
| SERPINB2 | 6.340134 | 5.738694 | 0.001279 | 1.104804 | 0.143791 | 0.014206 |
| COL4A2 | 126.1337 | 135.8197 | 0.001305 | 0.928685 | -0.10674 | 0.014206 |
| DKK2 | 3.452056 | 3.712034 | 0.00197 | 0.929963 | -0.10475 | 0.019308 |
| IGFBP1 | 4.473332 | 5.262762 | 0.004129 | 0.849997 | -0.23447 | 0.03679 |
| IFNG | 0.508028 | 0.480253 | 0.005681 | 1.057833 | 0.081112 | 0.046391 |
| COL4A1 | 120.7089 | 129.6466 | 0.007841 | 0.931061 | -0.10305 | 0.059106 |
| F13A1 | 33.13304 | 31.58468 | 0.014657 | 1.049023 | 0.069046 | 0.095758 |
| IL11 | 0.351676 | 0.368394 | 0.01427 | 0.954619 | -0.067 | 0.095758 |
| GUCY1A2 | 5.86866 | 6.169325 | 0.02413 | 0.951265 | -0.07208 | 0.147794 |
| SST | 17.46769 | 15.94985 | 0.033801 | 1.095163 | 0.131146 | 0.194852 |
| IGFALS | 1.412191 | 1.541967 | 0.035902 | 0.915837 | -0.12684 | 0.195464 |
| F11 | 5.20928 | 5.966597 | 0.041148 | 0.873074 | -0.19582 | 0.212236 |
| SERPINE1 | 30.60093 | 32.15194 | 0.045225 | 0.95176 | -0.07133 | 0.221602 |
| COL4A5 | 5.343238 | 5.511835 | 0.063907 | 0.969412 | -0.04482 | 0.272419 |
| ADCY1 | 0.934538 | 0.904675 | 0.063935 | 1.03301 | 0.046854 | 0.272419 |
| IFNA1 | 1.900172 | 1.645373 | 0.058477 | 1.154858 | 0.207716 | 0.272419 |
| IGFBP3 | 35.88824 | 37.81646 | 0.06762 | 0.949011 | -0.0755 | 0.276114 |
| F2R | 17.8615 | 18.57053 | 0.071365 | 0.96182 | -0.05616 | 0.27975 |
| GHRH | 0.133716 | 0.117468 | 0.075471 | 1.138313 | 0.186897 | 0.284467 |
| IL7 | 4.766534 | 4.597941 | 0.082626 | 1.036667 | 0.051953 | 0.299903 |
| HLA-DRB3 | 11.7147 | 12.42623 | 0.103898 | 0.94274 | -0.08507 | 0.363642 |
| IGFBP6 | 9.694764 | 9.706376 | 0.119517 | 0.998804 | -0.00173 | 0.390422 |
| DKK1 | 22.10085 | 22.91794 | 0.118374 | 0.964347 | -0.05238 | 0.390422 |
| NOS3 | 6.61765 | 6.748245 | 0.131233 | 0.980648 | -0.02819 | 0.403521 |
| TRPC4 | 2.317167 | 2.436718 | 0.134398 | 0.950938 | -0.07258 | 0.403521 |
| TRPC5 | 0.240457 | 0.234866 | 0.13588 | 1.023803 | 0.033938 | 0.403521 |
| IL3 | 0.040412 | 0.048445 | 0.153974 | 0.834189 | -0.26155 | 0.443809 |
| THY1 | 250.0147 | 256.8635 | 0.158908 | 0.973337 | -0.03899 | 0.444942 |
| KREMEN2 | 0.66296 | 0.714977 | 0.163883 | 0.927246 | -0.10898 | 0.446127 |
| CCR3 | 1.093817 | 1.05404 | 0.177757 | 1.037737 | 0.053441 | 0.470815 |
| HLA-DRA | 240.3654 | 232.0491 | 0.201984 | 1.035839 | 0.050799 | 0.520906 |
| FZD1 | 5.426793 | 5.19561 | 0.212446 | 1.044496 | 0.062807 | 0.533839 |

| | | | | | | |
|---|---|---|---|---|---|---|
| KCNQ2 | 0.616584 | 0.676582 | 0.217989 | 0.911322 | -0.13397 | 0.534073 |
| IGFBP5 | 139.5291 | 142.7088 | 0.253947 | 0.977719 | -0.03251 | 0.606994 |
| WNT8B | 0.196908 | 0.187153 | 0.266235 | 1.052122 | 0.073302 | 0.621216 |
| GHRL | 1.549805 | 1.969712 | 0.275682 | 0.786818 | -0.3459 | 0.628299 |
| IL1B | 9.159799 | 8.311716 | 0.28624 | 1.102035 | 0.14017 | 0.637535 |
| RAPGEF5 | 22.38004 | 23.16832 | 0.320002 | 0.965976 | -0.04994 | 0.640003 |
| PLAU | 13.92074 | 14.346 | 0.301644 | 0.970357 | -0.04341 | 0.640003 |
| CD3D | 10.69667 | 11.29104 | 0.302337 | 0.947359 | -0.07802 | 0.640003 |
| IGF1 | 5.536212 | 5.559563 | 0.308441 | 0.9958 | -0.00607 | 0.640003 |
| EPO | 0.189023 | 0.208045 | 0.319003 | 0.908568 | -0.13833 | 0.640003 |
| ITGB2 | 80.68734 | 76.83119 | 0.330846 | 1.05019 | 0.07065 | 0.648459 |
| ITGAX | 16.62007 | 15.83322 | 0.343258 | 1.049696 | 0.069971 | 0.659594 |
| IGFBP4 | 42.73864 | 43.66044 | 0.370802 | 0.978887 | -0.03079 | 0.661693 |
| ITGAL | 15.41324 | 15.15506 | 0.371358 | 1.017036 | 0.024371 | 0.661693 |
| COL4A6 | 0.729051 | 0.737712 | 0.369763 | 0.98826 | -0.01704 | 0.661693 |
| PLAT | 132.5589 | 130.0715 | 0.367849 | 1.019123 | 0.027329 | 0.661693 |
| CD8A | 3.415586 | 3.407509 | 0.404158 | 1.00237 | 0.003415 | 0.707277 |
| CCL11 | 2.96355 | 3.101497 | 0.424294 | 0.955523 | -0.06564 | 0.729488 |
| APP | 279.2316 | 283.0418 | 0.44122 | 0.986538 | -0.01955 | 0.74551 |
| CD28 | 3.54859 | 3.457023 | 0.480784 | 1.026487 | 0.037716 | 0.76157 |
| CD4 | 8.300176 | 8.062119 | 0.463251 | 1.029528 | 0.041983 | 0.76157 |
| IL2 | 0.266584 | 0.279494 | 0.479105 | 0.953809 | -0.06823 | 0.76157 |
| TLR7 | 2.848408 | 2.722911 | 0.482509 | 1.046089 | 0.065006 | 0.76157 |
| ANPEP | 55.2169 | 48.97576 | 0.489581 | 1.127433 | 0.173042 | 0.76157 |
| HLA-DRB5 | 33.96501 | 35.24904 | 0.510114 | 0.963573 | -0.05353 | 0.781112 |
| PTPRC | 0.345803 | 0.348093 | 0.55971 | 0.99342 | -0.00952 | 0.818681 |
| GNAS | 190.484 | 192.1411 | 0.555008 | 0.991376 | -0.0125 | 0.818681 |
| COL4A3 | 2.246328 | 2.306565 | 0.558027 | 0.973884 | -0.03818 | 0.818681 |
| CD5 | 3.249027 | 3.293036 | 0.579605 | 0.986636 | -0.01941 | 0.835313 |
| IL10 | 1.410248 | 1.284904 | 0.615607 | 1.097551 | 0.134288 | 0.86185 |
| HLA-DRB1 | 134.2702 | 131.7672 | 0.607662 | 1.018996 | 0.027148 | 0.86185 |
| CSF1 | 15.09522 | 14.63318 | 0.635041 | 1.031574 | 0.044848 | 0.876536 |
| COL4A4 | 1.664774 | 1.619377 | 0.645554 | 1.028033 | 0.039887 | 0.878671 |
| IL13 | 0.148691 | 0.166913 | 0.658406 | 0.890831 | -0.16678 | 0.883887 |
| CSF3 | 0.241441 | 0.270488 | 0.669021 | 0.892615 | -0.16389 | 0.886001 |
| CD3E | 3.611179 | 3.676768 | 0.679651 | 0.982161 | -0.02597 | 0.888077 |
| WNT8A | 0.212926 | 0.210431 | 0.699022 | 1.011857 | 0.017005 | 0.901371 |
| GUCY1B2 | 0.117637 | 0.114455 | 0.721064 | 1.027805 | 0.039567 | 0.916251 |
| IL5RA | 0.5349 | 0.525738 | 0.738611 | 1.017426 | 0.024924 | 0.916251 |
| CSF2 | 0.464305 | 0.490354 | 0.736615 | 0.946876 | -0.07875 | 0.916251 |
| KCNQ4 | 1.031623 | 1.061972 | 0.7602 | 0.971422 | -0.04183 | 0.925889 |

| | | | | | | |
|---|---|---|---|---|---|---|
| CD33 | 2.685025 | 2.571431 | 0.765276 | 1.044175 | 0.062364 | 0.925889 |
| CD40 | 6.996738 | 7.014987 | 0.785324 | 0.997399 | -0.00376 | 0.92725 |
| IFNB1 | 0.40215 | 0.398549 | 0.784699 | 1.009034 | 0.012974 | 0.92725 |
| KCNQ5 | 1.984083 | 2.036391 | 0.81366 | 0.974313 | -0.03754 | 0.938102 |
| IL5 | 0.918246 | 0.891812 | 0.81098 | 1.029641 | 0.042142 | 0.938102 |
| GH1 | 0.253986 | 0.240908 | 0.879246 | 1.054285 | 0.076265 | 0.94375 |
| CD247 | 1.340309 | 1.352437 | 0.894933 | 0.991033 | -0.013 | 0.94375 |
| CD3G | 2.645035 | 2.61187 | 0.898651 | 1.012698 | 0.018204 | 0.94375 |
| CD2 | 5.977274 | 5.972806 | 0.843204 | 1.000748 | 0.001079 | 0.94375 |
| TRPC3 | 0.60144 | 0.585058 | 0.876152 | 1.028 | 0.03984 | 0.94375 |
| ICAM1 | 17.30879 | 17.27843 | 0.863639 | 1.001757 | 0.002533 | 0.94375 |
| CD7 | 2.710846 | 2.767003 | 0.837295 | 0.979705 | -0.02958 | 0.94375 |
| TRPV4 | 9.437347 | 9.389004 | 0.905229 | 1.005149 | 0.007409 | 0.94375 |
| IL6 | 2.973981 | 2.991521 | 0.857324 | 0.994137 | -0.00848 | 0.94375 |
| IL9 | 0.120191 | 0.108006 | 0.944594 | 1.112816 | 0.154215 | 0.954332 |
| IL4 | 0.571577 | 0.553736 | 0.939582 | 1.032219 | 0.045749 | 0.954332 |
| KCNQ3 | 1.195197 | 1.204244 | 0.939356 | 0.992487 | -0.01088 | 0.954332 |
| IGFBP2 | 19.61493 | 20.58527 | 0.985116 | 0.952862 | -0.06966 | 0.985116 |

**Supplementary Table S2. List of differentially expressed genes between YOPC and AOPC.** Genes are listed in order of statistical significance (*Q*-value).
